# Clinical outcomes in hospitalized vaccine-breakthrough COVID-19 cases compared with contemporary unvaccinated hospitalized adults

**DOI:** 10.1101/2022.02.07.22270555

**Authors:** Marcelo Wolff, Margarita Gilabert, Rodrigo Hernández

**Author notes:** Corresponding author: Marcelo Wolff: Clínica Santa María, 410 Santa María Ave, Santiago, Chile.

## Abstract

**Summary:** The inactivated SARS-CoV-2 vaccine (CoronaVac®) has been the principal vaccine used in Chile’s pre-booster immunization campaign. We compared major outcomes in 260 hospitalized vaccinated vs 507 unvaccinated adults with COVID-19 (mid-2021). The vaccinated group was much older, required less critical care, had lower hospital mortality (adjusted by age), and had shorter hospitalization than the unvaccinated. Benefits were most pronounced in those older than 59 years

**Objective:** To compare major outcomes in fully vaccinated and unvaccinated adult persons hospitalized for COVID-19 in a general private hospital in Santiago, Chile during mid2021.

## Background

The present array of SARS-CoV-2 vaccines have variable efficacy in clinical trials and effectiveness in national “real-world” vaccination programs. Their preventive performance can be evaluated by avoidance of symptomatic infection and severity of disease, the latter often inferred from required level of care (hospitalization, critical care unit [CCU] admission, ventilator use) or mortality. Messenger RNA (mRNA)-based vaccines have shown the most potent preventive effect for both the ancestral SARS-CoV-2 strain and the subsequent variants like delta and omicron (1,2). Nonetheless, breakthrough cases occur since vaccines do not confer sterilizing immunity; cases following mRNA-based vaccines are far less likely to be severe.

Chile has implemented an early and expansive vaccination program that enabled two-dose immunization in 88% of its 19.5 million population, older children included, by end of the third quarter of 2021 (3). CoronaVac® (Sinovac Biotech Ltd, Beijing, China) is an inactivated vaccine that has been the cornerstone of the program, used in 75% of vaccinated adults. BNT162b2 (Pfizer with BioNTech and Fosun Pharma, New York, NY, USA) mRNA vaccine has been used mostly in immunocompromised adults and minors.

CoronaVac immunogenicity is comparatively less robust, though substantial antibody levels are achieved; real-world effectiveness was assessed in a Chilean study of 4 million vaccinees demonstrating a 65.9% (95% CI 65.2-66.6) preventive effect for symptomatic infection and much higher prevention of hospitalization, 87.5% (95% C.I. 86.7-88.2), critical care units (CCU) admissions, 90.3% (95% C.I. 89.1-91.4), and death, 86.3% (95% C.I. 84.5-87.9) in the short run (4). Effectiveness declined with time (5), suggesting the benefit of boosting doses.

## Methods

We reviewed clinical and demographic information of disease characteristics in two groups: (1) fully vaccinated (at least 14 days post-second dose, but not boosted), and (2) non-vaccinated adults (> 17 years) admitted for COVID-19 in a 353-bed private hospital. We used data extracted from a disease-specific database created at the beginning of the COVID-19 pandemic and enhanced data with relevant information incorporated in real time. Data were extracted from March 27, 2021 (the day the first fully vaccinated case was admitted) to August 31, 2021; this Southern Hemisphere winter season coincided with the second wave of COVID-19 in Chile. Data included: age, sex, CCU admission, use of mechanical ventilation, length of hospitalization, and intra-hospital death.

A case of COVID-19 was defined by a positive PCR or characteristic clinical and radiologic findings. Accurate information on the type, number, and date of vaccination doses was provided from a mandatory nationwide registry. Our local ethics committee authorized use of these data for outcomes research and waived the need for informed consent, under conditions guaranteeing data confidentiality and patient anonymity. Quantitative variables were compared by Student’s t test and categorical variables were analyzed with proportional Z tests using Stata® 13 (StataCorp LLC, College Station, TX, USA) software. Comorbidities at time of admission were not evaluated and no genetic characterization of SARS-CoV-2 strains was performed.

## Results

During the study period there were 998 adult COVID-19 admissions (2.5% in the 2 comparative groups with undocumented positive PCR, excluding partially vaccinates cases), 260 in fully vaccinated [Fvac] (26.1%), 507 in unvaccinated [Unvac] (50.8%) and 231 with incomplete vaccination either by dose or timing (23.1%). Data for persons with incomplete vaccination were excluded from the study. Vaccines courses were overwhelmingly CoronaVac (251 or 96.5%), with the rest BNT162b2 (3.1%); Convidicea (Ad5-nCoV; CanSinoBIO, Tianjin, China; 0.4%). Median age (IQR_25-75_) for Fvac was 64.8 (57-75) years and for Unvac 45 (35-53) years (p < 0.001). Fvac hospitalized patients were more likely to be older than 60 years of age than Unvac patients (69% vs. 15.6%, p <0.001). Despite the older age, Fvac admission to the CCU was less common (33.8%) than for Unvac (43.1%; p=0.012). Similarly, the need of mechanical ventilation was 13.8% for Fvac and 26.4% for Unvac (p <0.001). Unadjusted for age, there were 45 deaths in Fvac (17.3%) and 65 in Unvac (13.4%) groups (p=0.15). A stratified analysis of death by decade of age is informative (Table). Total mean (and median) length of stay in days were 11.7 (9) of Fvac and 12.6 (10) for Unvac; for those age ≥60 years Fvac length of stay was 12.8 (10) days and 17.8 (15) days for Unvac (p < 0.001; Table).

**Table.** Comparative data in hospitalized COVID-19 patients in Chile with and without full SARS-CoV-2 vaccination prior to diagnosis.

|  | Fully vaccinated | Unvaccinated | P value |
| --- | --- | --- | --- |
| Enrollment: N (% of COVID-19 admissions) | 260 (26.1%) | 507 (50.7%)* |  |
| Mean age in years (median) and [IQR<sub>25-75</sub>] | 64.8 (67)<br>[57-75] | 45.5 (45)<br>[35-53] | <0.001 |
| Persons >60 years | 69.0% | 15.6% | <0.001 |
| Women | 36.9% | 39.6% | 0.46 |
| CCU admission | 33.8% | 43.1% | 0.012 |
| Mechanical ventilation | 13.8% | 26.4% | <0.001 |
| Total mortality | 45 (17.3%) | 68 (13.4%) | 0.15 |
| ≥80 years | 13/38 (34.2%) | 2/9 (22.2%) | 0.49 |
| 70-79 | 12/69 (17.4%) | 6/21 (28.6%) | 0.26 |
| 60-69 | 15/72 (20.8%) | 16/44<br>(36.3%) | 0.066 |
| 50-59 | 3/39 (7.7%) | 19/104<br>(18.3%) | 0.12 |
| 40-49 | 1/21 (4,8%) | 18/141<br>(12.8%) | 0.29 |
| 30-39 | 1/17 (5.9%) | 7/134 (5,2%) | 0.91 |
| <30 | 0/4 (0) | 0/54 (0) | - |
| ≥60 | 40/179 (22.3%) | 24/74<br>(32.4%) | <0.001 |
| Mean and (median) length of hospitalization<br>(days) | 11.7 (9) | 12.6 (10) | 0.35 |
| ≥60 years | 12.8 (10) | 17.8 (15) | <0.001 |

\*= There were 231 (23.2%) COVID-19 admissions in adults with incomplete vaccination and/or <14 days of last dose at COVID-19 diagnosis; these patients were not eligible for the study
CCU: Critical Care Unit

## Discussion

This study of adults hospitalized for COVID-19 during the second wave in Chile found that hospitalizations, CCU admissions, and ventilator use were all more common in unvaccinated people at a time when the majority of the adult population had been vaccinated with inactivated SARS-CoV-2 vaccines. When age is considered, deaths are also lower with immunization. The U.S. Centers for Disease Control and Prevention has reported similar results with mRNA vaccine use (6). We believe that the good performance of the less potent inactivated vaccine is important for China and the >45 low- and middle-income countries (LMIC) that use World Health Organization-approved inactivated vaccines as the backbone of their vaccine efforts (7). Chilean data have confirmed inactivated vaccine efficacy in clinical trials (8) and effectiveness for multiple outcomes in a state sponsored vaccination programs in > 4 million people (4). Our study demonstrates the value of inactivated vaccines in reducing severity of disease in hospitalized patients with COVID-19 at multiple age strata. Among the hospitalized COVID-19 cases, one in four had received full primary vaccination, overwhelmingly CoronaVac (96.5%), much higher than its 75% participation in the mix of vaccines used in the country consistent with the much higher hospitalization prevention effect found with BNT162b2 in the country compared with the former during the same period of this study (97.2% vs 86%) (5).

Our vaccinated hospitalized subjects were two decades older, on average, than the unvaccinated hospitalized subjects, almost certainly representing a population with more baseline comorbidities and naturally more prone to have severe disease and death when infected compared to younger people. The higher mortality in the vaccinated group as a whole is misleading; once age is controlled for by stratum-specific analyses, death rates in each decade group from the 40s to the 70s were lower in vaccinees, with too few cases ≥ 80 years in the unvaccinated group for meaningful comparisons. This older cohort also had fewer admissions to CCUs, less need for mechanical ventilation and shorter lengths of hospital stay as a whole; these differences were most pronounced and statistically significant in those older than 59 years of age. These clinical outcomes data suggest that full prior vaccination with the inactivated vaccine provides a significant protection from serious disease. At a national level, the aggressive vaccination program in Chile of early 2021 seems to have blunted the magnitude of the epidemic, including the more salutary consequences for those with breakthrough disease, reinforcing confidence in the vaccines used. We will be unable to follow-up this study since unvaccinated persons are now scarce with special incentives towards vaccination and restrictions on the unvaccinated being put in place for adults and children in a national effort to maximize immunization in the population. A vigorous booster vaccination campaign for all with fully primary vaccination, beginning with the elderly, has been conducted in late 2021 and 2022. While there is a scarcity of unvaccinated people, we intend to compare COVID-19 cases in vaccinated people with and without booster doses. Limitations of our study include a lack of evaluation of comorbidities and viral variants, being a single-site study, risk of patient’s profile not being an accurate representation of the COVID-19 population at large and portraying a situation prior to the Omicron surge. Our design does not evaluate the decline of preventive effects of the CoronaVac over time. However, we believe that Chilean data are reassuring to China and LMICs since there are many countries with vaccination programs in their infancy that are or have been heavily dependent on inactivated vaccines. The global community may wish to consider mRNA boosters for persons with primary vaccination with inactivated vaccines, given highly favorable immunological results of this combination noted from the Dominican Republic (9) and enhanced preventive effectiveness from Chile (10).

## Data Availability

Data used for this work is embedded in a authorized institutional database especific for COVID-19 hospitalized patients in the institution, only accessible upon authorization

## Notes

### Author contributions

MW and RH contributed to the concept and design of the study. MW and MG contributed to acquisition of data. MW RH and MG contributed to interpretation of data. All authors contributed to drafting or revision of the manuscript. MW and RH contributed supervision for the study. Acknowledgments. We acknowledge the invaluable assistance of Andrea Canals for statistical analysis. No external or specific funds were required for this study

